# Correlation between meteorological factors and COVID-19 infection in the Belém Metropolitan Region

**DOI:** 10.1101/2020.06.10.20127506

**Authors:** Félix Lélis da Silva, Maryjane Diniz A. Gomes, Andréa P. Lélis da Silva, Samio Costa de Sousa, Marcos Francisco Serafim de Souza, Gabriel Lélis P. da Silva

**Author notes:** **Corresponding author** (s): Félix Lélis da Silva.

## Abstract

Many factors can influence the spread of viruses and respiratory infections. Studies have suggested that there is a direct relationship between environmental issues and population density with cases of COVID-19. In this sense, this research aims to analyze, through correlational study and Krigagem, the relationship of meteorological and demographic variables with cases of COVID-19 in regions of subtropical climate in Brazil. The results suggest that population and demographic density (hab/km^2^) are risk factors for the spread of SAR-Cov-2 and an increase in the daily case record of COVID-19. The distribution of cases according to age group did not present a significant disparity between men and women. Relative humidity (RH)%, average temperature °C, minimum temperature °C, maximum temperature °C, wind speed m/s and daily precipitation (rain) mm show negative relationships with cases of COVID-19 in regions of humid equatorial climate. Analysis between associations of environmental factors, wind, temperature and HR in a region is extremely important to understand the dynamics of SARS-Cov-2 in the environment. In the northern region of Brazil, low wind speed, high temperatures and high RH are observed, environmental factors that, when associated, reduce the transmission process because it hinders the movement of the virus in the environment. In this sense, it is suggested that the transmission of SARS-CoV-2 in this region is disseminated through fluids in the air between man/man and by contact between objects/men. Therefore, strategic public policies to combat the pandemic must consider the environmental factors of the regions involved and control and/or blocking the transit of people.

## 1. Introduction

The outbreak of the disease caused by the new coronavirus (COVID-19) institutes a worldwide public health emergency and, due to its high spread, internationally, a pandemic has been officially declared by the World Health Organization (WHO, 2020). COVID-19 is a viral infection that causes a severe acute respiratory syndrome and can cause progressive respiratory failure and death (Coccia, 2020).

Wuhan, China, was considered the first metropolitan region to be the center of the epidemic in December 2019 (Zhou et al., 2020; Wang et al., 2020). In the region, studies infer that several environmental factors contributed to the spread of the infection (Bhattachaejee, 2020; Liu et al., 2020; Ma et al., 2020).

The transmission factors of a viral disease can be as variable as possible. The spread of a virus is associated with environmental factors (temperature, wind speed and relative humidity), population density and organization, efficiency of the health sector, and general factors, such as the biological characteristics of the virus, time of incubation, effects on infected and susceptible people (Wang et al., 2020).

Meteorological factors play an important role in the spread of SARS Cov-2. In China, a country with climatic diversity, low temperature and relative humidity ranges favored the transmission of the virus (Liu et al., 2020). The transmission rate of COVID-19 has low sensitivity to changes in ambient temperature and high sensitivity to population size (Jahangiri, Jahangiri, Najafgholipour, 2020).

Studies have suggested that temperature is a conductor of COVID-19 (Shi et al., (2020a), and acts as an indicator of transmission of SARS-COV-2. According to Tan et al. (2005), the incidence of COVID-19 presents different dynamics in temperature variations in the region, so to analyze the spread of the SARS-COV-2 virus associated with environmental factors, one must consider the climatic and demographic characteristics of the region where the infection occurred.

Although 90% of COVID-19 cases have been reported in countries with a temperature range of 3 to 17 ° C (Bukhari and Jameel, 2020), SARS-CoV-2 can survive for several days in plastics and metals at temperatures moderate (between 21 and 23 ° C) and an RH of 40% (Van Doremalen et al., 2020). This suggests that there may be a direct relationship between temperature, humidity, air pollutants, the environment and the spread of SARS-CoV-2.

Lal et al (2020) state that there is no evidence to support that cases of COVID-19 reduce in hot climates. However, studies suggest that temperature in tropical regions has an influence on case reports of influenza (Tamerius et al., 2013). Likewise, environmental factors have an influence on the dynamics of the spatial distribution of infectious diseases (Lemaitre et al., 2019). These results are useful, as they provide implications for public policy makers in regions with different climatic conditions.

It is necessary to conduct studies in other regions, during the pandemic, to assess the implication of these environmental factors in the transmission of COVID-19. Researchers estimate that climatic variables explain 18% of the variation in the time of disease transmission and 82% are related to containment measures, health policies, population density, transport or cultural aspects (Oliveiros et al., 2020).

In the Americas, the number of infected people has evolved rapidly, with more than 1.5 million cases and 50 thousand deaths in the period of this study (PAHO, 2020). In the Americas region, Brazil stands out as the second country in number of incidence and mortality (WHO, 2020).

According to Hotez et al. (2020), regions such as Brazil, Mexico, Pakistan, Philippines and South Africa are vulnerable to the spread of SARS-CoV-2. The researchers suggest the possibility of an unprecedented advance in infections and mortality in poorer regions in tropical and subtropical countries.

Brazil has a great climatic variety and presents regions with high demographic densities associated with low levels of human development. In Latin America, Brazil has shown an increase in the registration of cases and deaths due to COVID-19, to the point of being classified as a new epidemic center for SARS-CoV-2 by the World Health Organization.

1. Do the cases of COVID-19 present a gender/gender gap in the BMR?
2. Are meteorological variables risk factors for the advance of the pandemic in the region of the Pará State, Northern Brazil?
3. Which environmental factors are correlated to 5% significance with cases of COVID-19 in the region?
4. Can high temperatures and RH in the region favor the reduction of the SARS-Cov-2 outbreak and the registration of new cases of COVID-19?
5. Can the beginning of the less rainy period in the Amazon region contribute to the reduction of COVID-19 cases?
6. What are the considerations for regions with similar climatic behavio?

## 2. Materials e methods

An epidemiological and correlation analysis was performed on the demographic characteristics of the diagnosed infected. The variables evaluated were: sex/gender, age group, population and demographic density. Subsequently, the correlation between environmental variables such as, average temperature °C, minimum temperature °C, maximum temperature °C, Relative Humidity % (RH), precipitation (mm) and wind speed (m/s) (INMET, 2020) and the registration of new daily cases of COVID-19 (BRASIL, 2020).

### 2.1. Study area

The present epidemiological study was carried out in diagnosed cases of COVID-19 in the seven municipalities that make up the Belém Metropolitan Region (BMR), Pará (Fig.1).

BMR has a humid equatorial climate, a high population record, with about 2,275,032 inhabitants and varied demographic density. Its municipal regions have a subtropical climate with wide variations in temperature and high rainfall, distributed over two seasons (more and less rainy). The months between June and December are considered the rainiest months in the Amazon (SOUZA FILHO et al., 2005; MORAES et al., 2005).

BMR is composed of 7 municipalities, they are: Belém, Ananindeua, Marituba, Benevides, Castanhal, Santa Barbara do Pará and Santa Izabel do Pará (IBGE, 2020). During the study period the Relative Humidity (%) in the regions is distributed between 80 to 100% with an average of 90.3%. The temperature range was 24.7 to 34.8 °C, daily rainfall was 0 to 97.8 mm and the recorded wind speed was between 0 to 3,601 m/s (INMET, 2020).

### 2.2. Data collect

The collection period ranged from March 18, 2020 to May 6, 2020, a period that preceded the implementation of the “Lockdown Regime”. Data on COVID-19 cases registered in the Belém Metropolitan Region (BMR) were obtained from the website of the Ministry of Health of Brazil (Brazil, 2020) and Health Department of the Pará State (SESPA, 2020). Meteorological data were obtained from stations located in the regions of Belém, Cametá, Breves, Castanhal, Tracuateua and Tucurui (INMET, 2020).

Figure. 2 shows the epidemiological situation (on May 5, 2020) in the Metropolitan Region of Belém (RMB). Although the first COVID-19 case was announced in the world in late December 2019 (şahin, 2020; Zhou et al., 2020), the first case in Brazil was reported on February 26, 2020. In the city of Belém, state of Pará, northern Brazil the first case, a 37-year-old man, was notified on March 18, 2020, exactly 23 days after the first case registered in the country (SESPA, 2020).

**Fig 1.**
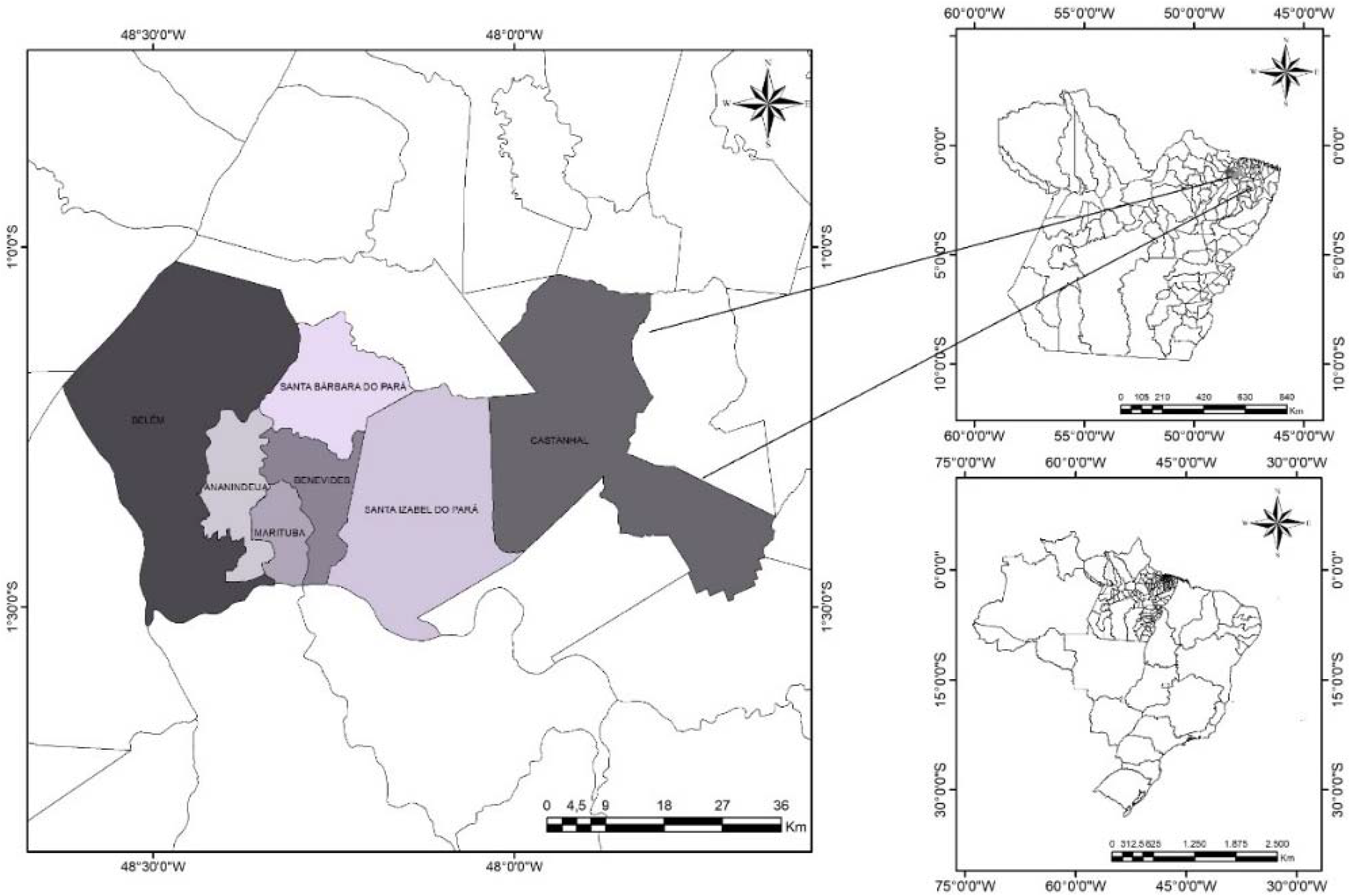
Composition of the Belém Metropolitan Region - BMR

**Fig 2.**
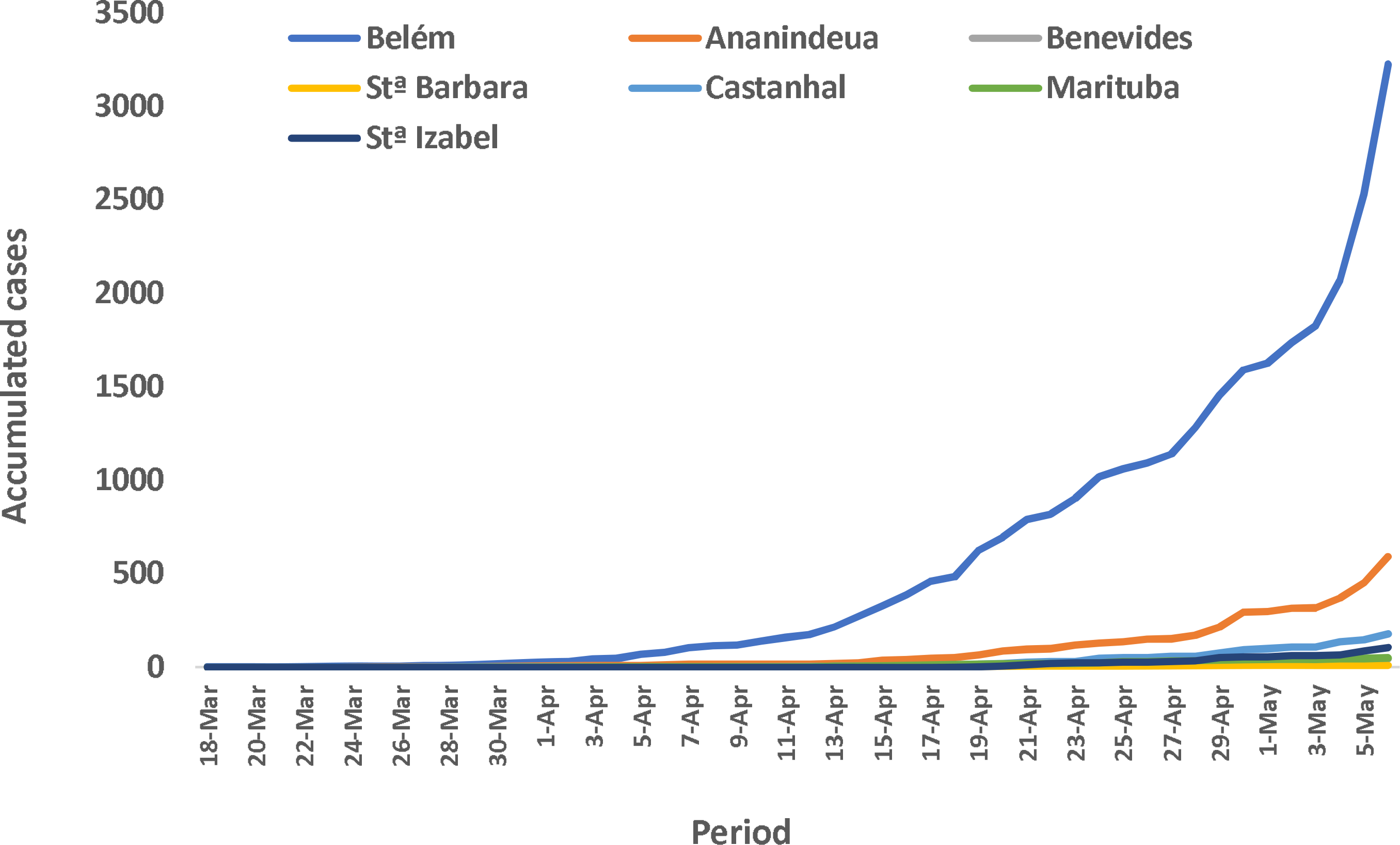
Case records of COVID-19 accumulated in the municipalities of BMR

### 2.3. Statistical analysis

The descriptive analyzes of the variables were expressed as mean and number (%). Cases without identification for age and gender were imputed. The proportions and mean between gender were analyzed using the Chi-square test of adherence and the Independence T test considering homogeneity of variance of the data with a significance alpha level 5%.

The distribution of cases within genders was assessed using boxplot diagrams. The occurrences of cases and the mean age of the infected were analyzed using the Chi-square test and the Independence T test. The correlations between number of cases and environmental variables were analyzed using the Spearman test.

Spearman’s nonparametric test was applied due to the monotonic relationship between ordinal variables and non-compliance with the assumption of normality. It was used to assess the association between the number of COVID-19 cases and environmental factors. Spearman’s coefficient (Eq.1) was calculated according to (Sahin, 2020).

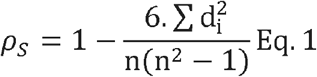

Where: *ρ*_*S*_ is the Spearman coefficient, *n* the number of alternatives and *di* represents the difference between the rows of the corresponding variables.

For the interpretation of the results, the magnitude of the correlations followed the following classification: correlation coefficient rho <0.4 implies a weak correlation, between 0.4 ≤ rho <0.5 moderate correlation and rho ≥ 0.5 a strong correlation between the variables evaluated (Hulley et al., 2003). The maps of temperature, relative humidity, precipitation and number of cases were structured using Krigagem spatial interpolation algorithm.

## 3. Results and discussions

In this section are presented: descriptive epidemiological and demographic analysis, proportional analysis (Chi-square test) for the occurrence of cases between sex/gender and parametric analysis (t test) to assess the age factor within the sex/gender of those infected. Next, a simple and multiple correlational analysis between meteorological variables and COVID-19 cases is demonstrated.

### 3.1. Epidemiological and demographic analyzes

In the period from March 18, 2020 to May 6, 2020, about 3969 cases were reported in the BMR by the State Health Department of Pará. Of these, 1986 (51.04%) were women and 1905 (48.96%) men. The average age of men was 48.11 ± 17.94 years with relative variability of CV = 37.29% and of women 46.51 ± 17.45 years and CV of 37.53%. Of these, 16 cases presented with no notification regarding sex/gender and 62 due to lack of identification regarding the age of the infected person.

As for the occurrence of cases by gender/sex in the municipalities of BMR, women were the most affected in Ananindeua (54.9%), Belém (51.2%) and Santa Izabel (51.5%) of the cases registered in the study period. Opposite results were obtained by Nikpouraghdam et al. (2020) for (sex/gender), with most infections registered in 66% of men and 44% in women. However, the cases recorded in men were proportionally higher in the municipalities of Marituba (67.4%), Benevides (63.6%), Castanhal (60.9%) and Santa Barbará (55.6%).

Table 1 lists the number of cases of COVID-19 in the municipal regions that occurred in the period. Epidemiological results indicate that most records occurred in the state capital Belém with 3223 (76.87%) cases, followed by Ananindeua (591; 14.09%), Castanhal (176; 4.20%), Santa Izabel (103; 2.46%), Marituba (47; 1.12%), Benevides (44; 1.05%) and Santa Barbara (9; 0.21%).

**Table 1.**
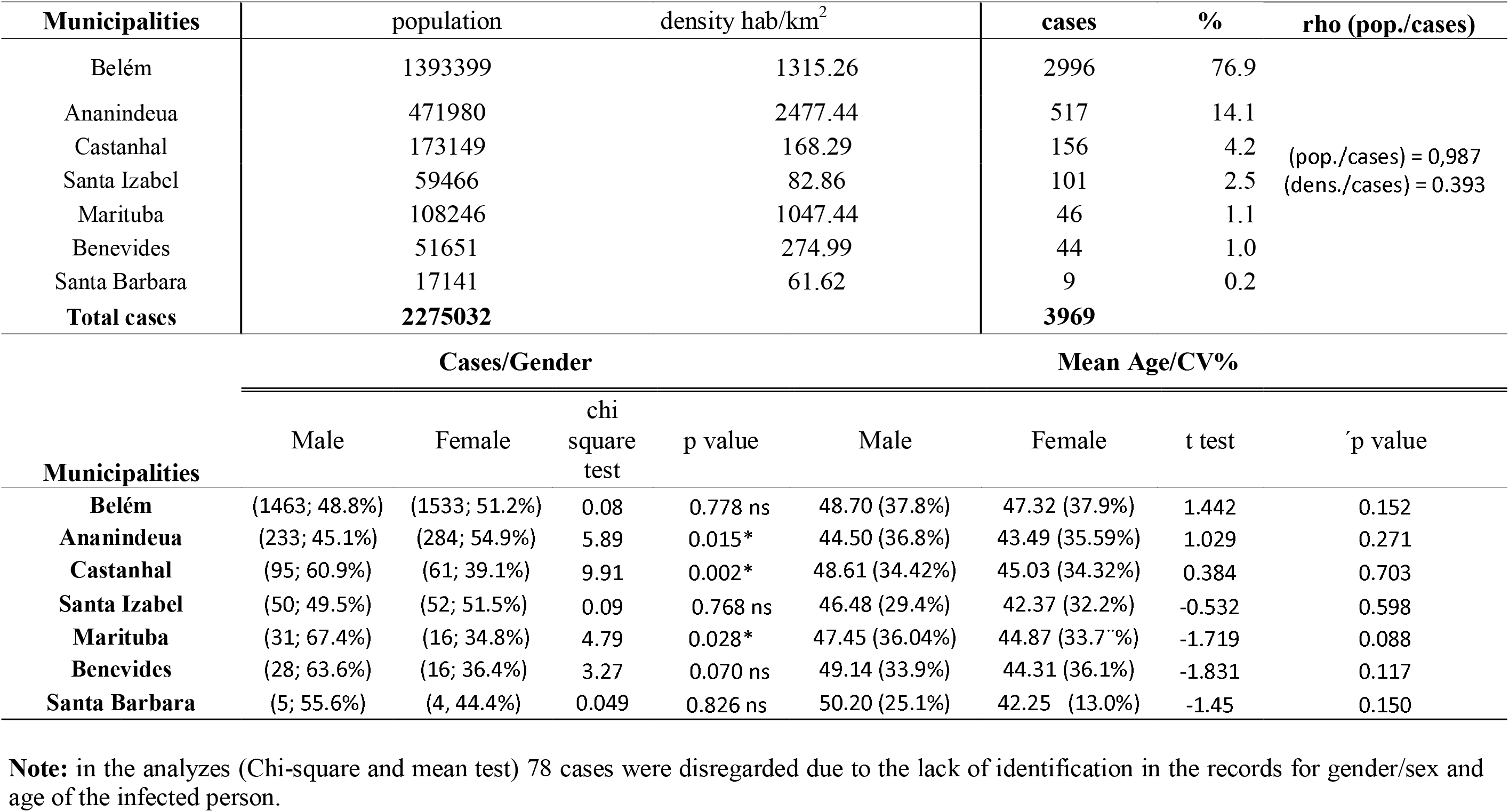
Relational behavior of demographics and new cases of COVID-19 recorded in Belém Metropolitan Region.

The capital Belém has the largest population (1,393,399 inhabitants) and a population density of 1,313, .26 inhabitants per square kilometer. The region was the dissemination center for the SARS-CoV-2 virus, through community transmission, to neighboring municipalities that comprise the BMR, as well as to other regions of the state.

The city of Belém had the highest number of infected people among the seven municipalities that make up the BMR with 2996 cases (65.9‥%), followed by Ananindeua (517; 14.1%) and Castanhal (156; 4.2%) of the records in the period. It is worth mentioning that these municipalities are located on the margins of the BR Highway 316, which connects Capital Belém to other cities in the country and to several other municipalities of Pará State.

### 3.2. Non-parametric and parametric analyzes

In Table 1, the Spearrman test (rho = 0.987) was applied to assess the relationship between population and density in the occurrence of cases of COVID-19. The results show that there is a high positive correlation between the number of registered cases and the total population of these regions. However, demographic density and cases of infection by COVID-19 with (rho = 0.393) show a moderate correlation. Similar results were obtained by Sahin (2020), considering that urban centers with large agglomerations are closely linked to high infections by COVID-19.

The Chi-square test to assess the existence of a significant difference between cases occurring by gender in the municipalities in the region (Table 1) demonstrated a significant proportional difference of 5% between those infected by gender (male and female) in Ananindeua, Castanhal and Marituba. In the other regions, no proportional difference was observed, results reinforced by Figure 3A. These results show that sex/gender does not depend on the number of registered cases of SARS-CoV-2 infections. Therefore, sex is not associated with a risk factor for COVID-19.

**Fig 3.**
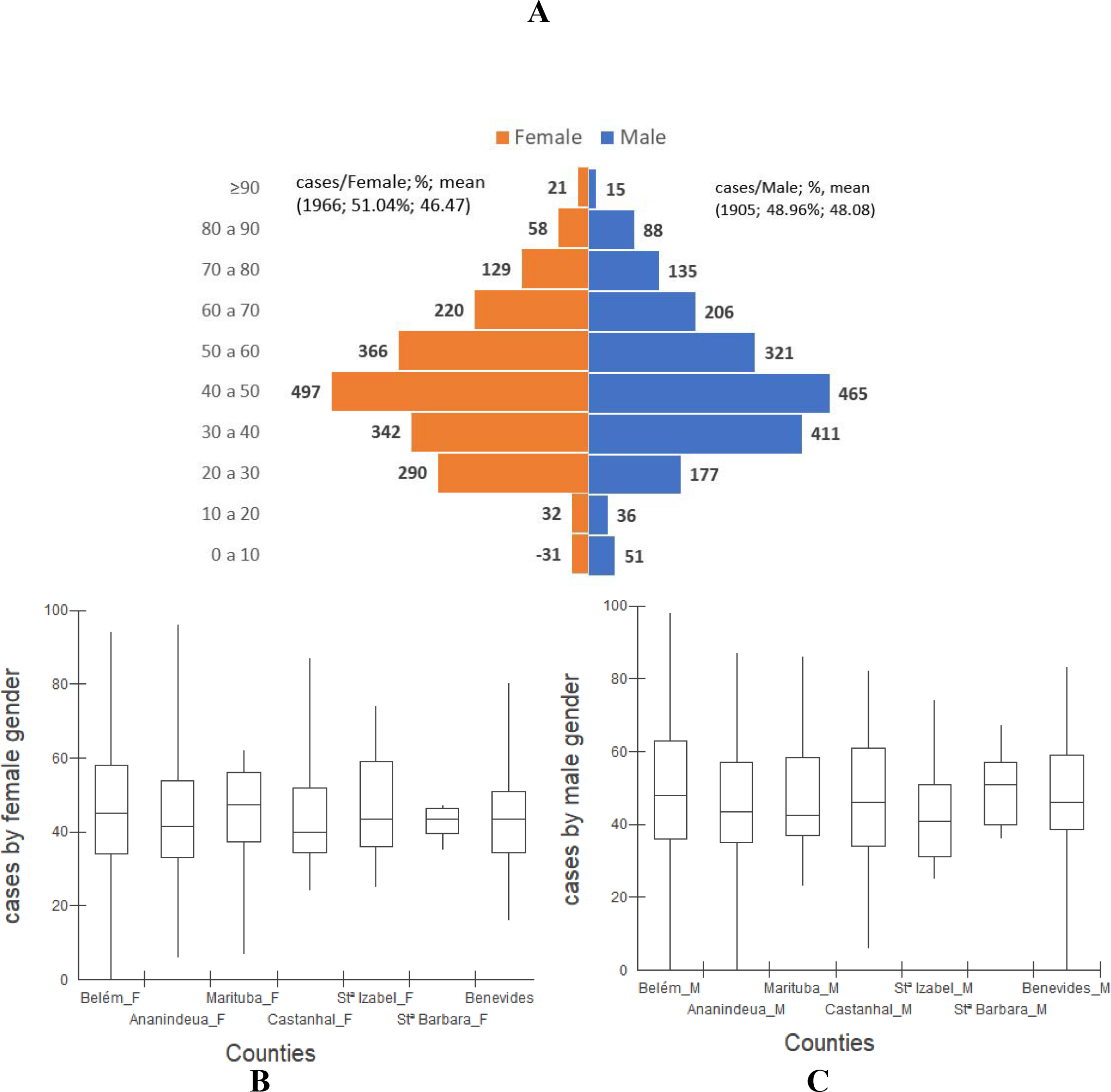
**A**) Behavior of the distribution of the total cases of COVID-19 according to age group and **B, C**) distribution by gender of the infected diagnosed in the municipalities of BMR

The Independence T tests, applied to compare the average ages of those affected by the virus by gender within the municipalities, were not significant at 5%. These tests demonstrate that there are no significant differences between the mean ages and that there is no difference in the behavior of the distribution of cases by age group between men and women.

This fact can be proven through the similarity of the variation coefficients (CV%) of the ages (Table 1) and Figures 3 B and C. These results show that within the municipal regions, which make up the BMR, the age group factor is not the COVID-19 is a risk factor. However, the economically active population aged 20 to 60 years is the most affected by the disease.

### 3.3. Simple correlational analysis between meteorological factors and COVID-19

Meteorological variables and their parameters are among the main causes that affect the dynamics of the spread of infectious diseases and the spread of viruses. Analyzing Spearman’s correlation coefficients with the respective levels of probability of the test (p-value), it is observed that environmental factors such as RH, average temperature, minimum temperature, maximum temperature, precipitation and wind, present a negative correlation with the numbers of COVID-19 cases in the municipalities that make up the metropolitan region of Belém (Table 2).

**Table 2.**
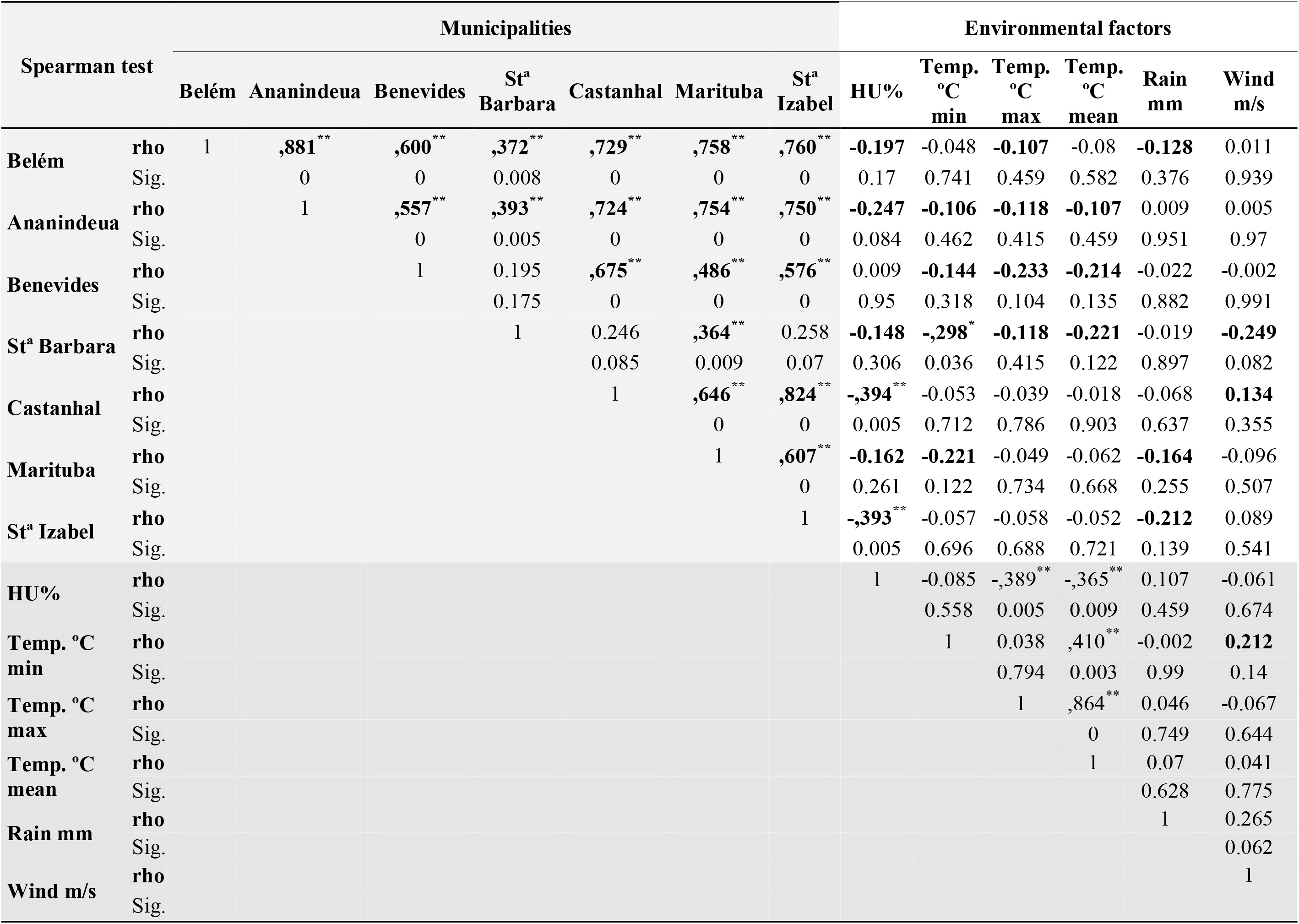
Spearman test to assess the correlations between environmental variables and COVID-19 case records, in the cities of BMR.

RH presented a negative, but not significant, correlation with the number of infection cases in the municipalities of Belém, Ananindeua and Santa Barbara. However, the number of COVID-19 cases did not correlate with RH in the municipality of Benevides. The region of Santa Izabel and Castanhal showed a significant negative correlation at 1% (p = 0.005) with the daily record of infections. Negative correlations for cases of COVID-19 and RH % were obtained by Sahin (2020) in Turkey, contributing to the present study.

This pointed out that the case/day records of infected people in these municipalities are significantly associated with RH. The records also demonstrate that an increase in RH implies a reduction in cases of COVID-19 in these locations. Likewise, studies carried out by Chan et al., (2020), suggest that high RH eliminates the viability of the SARS-CoV-2 virus, corroborating the results presented.

It is worth noting that the environmental factor RH%, in the region of the study in the evaluated period, is positively correlated to the rain factor and negatively correlated with average temperature and its maximum variation.

The minimum temperature recorded in the region has a negative correlation with the number of cases of COVID-19 recorded in the municipalities of Ananindeua, Benevides, Santa Barbara and Marituba. Temperature, despite the weak magnitude of correlation, stood out in the Santa Barbara region; that is, in this location, the minimum temperature in °C is associated with statistical significance to the number of daily infection case records.

Similar results, when assessing the spread of the number of cases of the SARS virus in the region of Beijing and Hong Kong to Bi et al., (2007) and in the region of Turkey according to (Sahin, 2020), concluded that the temperature is closely related, because at high temperatures a reduction in case records was observed.

The average temperatures recorded in the municipalities of Ananindeua, Benevides and Santa Barbara show a negative correlation to the cases of COVID-19 recorded in these regions. However, it has been proven through Spearman’s test that there is no evidence to confirm the significance of such relationships.

Bashir et al., (2020), after studies of the patterns of climate change in New York on the expansion of COVID-19, suggest that the average and minimum temperatures in the region were correlated with the case record. Results consistent with those observed in this study for BMR were also observed by Shi et al. (2020b), in mainland China. For, the researchers identified negative correlations of temperatures and Relative Humidity in the cases of COVID-19.

The maximum temperature °C showed a negative correlation with the number of cases of SARS-CoV-2 infections in the municipalities of Belém, Ananindeua, Benevides and Santa Barbara. However, without statistical evidence to suggest that this correlation occurs significantly.

Results obtained by Prata, Rodrigues and Bermejo (2020), for Brazilian cities, indicated the same negative behavior for linear correlation between temperatures and numbers of confirmed cases. However, studies by Shi et al. (2020) identified a correlation between the increase in the number of COVID-19 cases at temperatures above 10°C.

According to Chatziprodromidou et al., (2020) high temperatures and high humidity reduce the transmission of COVID-19. Tosepu et al., (2020), suggest that the climate is an important factor in determining the incidence rate of COVID-19, and the temperature and its variations are correlated with the registration of infections.

Regarding the environmental factor rainfall, the results show that there is a negative correlation with the number of cases of infections in Belém municipalities, Santa Barbara and Castanhal. However, this association is not statistically significant. Equivalent results of correlation of precipitation with cases of covid-19 were identified by Menebo (2020).

Wind speed in the cities of Santa Barbara showed a negative correlation with daily records of cases of COVID-19. However, in the Castanhal region during the study period, wind speed was positively correlated with cases of COVID-19. This leads us to believe that the higher the wind speed in the region, the greater the spread of the SARS-CoV-2 virus. Although these results were not significant at 5%, using the Spearma rho test, it is very important to highlight that, results obtained by Sahin (2020) in Turkey, demonstrate a positive correlation between cases of COVID-19 and the meteorological factor speed of wind in m/s of the region.

### 3.3 Multiple correlational analysis between meteorological factors and COVID-19 cases

For multiple associations between temperature variations (minimum and maximum) and cases of COVID-19, identified daily, it is noted that with temperatures between 24 to 34°C, minimum temperatures in the 24°C range, associated with RH between 85 to 95% and temperatures around 24°C and speed between 0.5 to 2.0 m/s, the lowest average daily records of COVID-19 were observed in the municipality of Santa Barbara (Fig.4).

These results suggest that regions of low density hab/km^2^ have lower records of COVID-19 cases. However, urban centers with a population density of more than 150 inhabitants per square kilometer recorded the highest number of infections in the region.

In Castanhal and Santa Izabel, significant negative correlations were found between the number of cases and relative humidity (Table 2). Multiple spatial correlation analysis was performed considering RH as the main factor for these regions. There are associations between RH/wind speed and RH/rain, suggesting that when the RH is between 85 to 100% and the wind speed in the range of 0 to 1 m/s, in Castanhal, smaller cases were registered of COVID-19. However, for higher winds of 2 m/s and daily rainfall above 60 mm, the highest numbers of infected cases were identified.

Similar results, from lower occurrences of COVID-19 cases, were found in Santa Izabel when the RH varied between 85 to 100% and rainfall levels from 0 to 30 mm average/day.

In Belém city, the combination of RH between 80 and 95% and rainfall between 0 and 30 mm/day was the lowest number of cases of SARS-CoV-2 infections. For RH less than 85% and daily rainfall greater than 60 mm favored the number of cases of COVID-19. Combination of RH in the 80 to 95% range, with an average temperature index between 26 and 32 °C, the lowest numbers of SARS-CoV-2 infected individuals were registered in the municipality of Ananindeua (Fig. 4).

**Fig 4.**
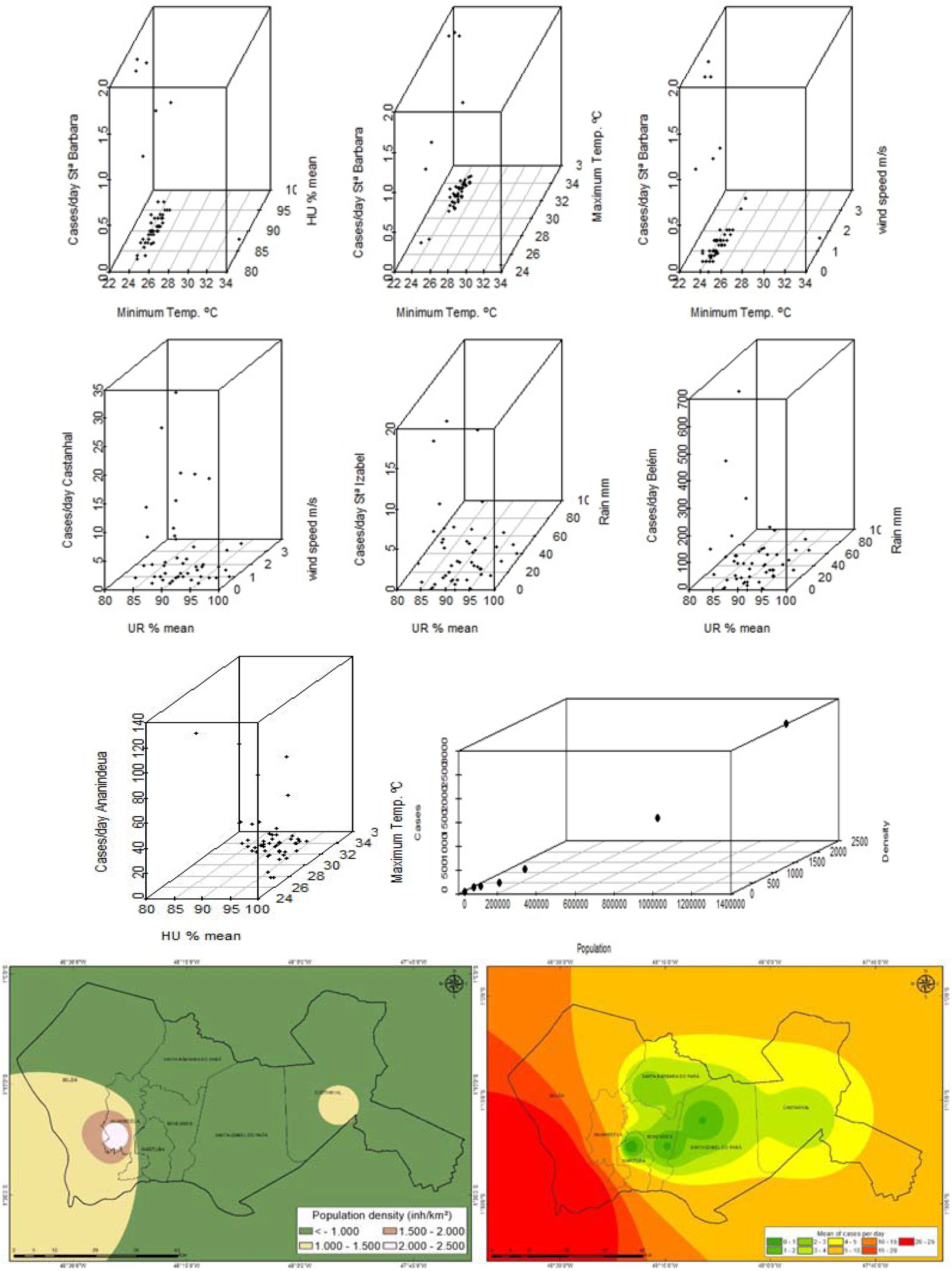
Multiple correlation between meteorological and demographic variables in the cases of COVID-19 in the BMR municipalities

Many factors can influence the spread of viruses and respiratory infections. Correlational studies aim to subsidize intervention measures, as they have implications for the structuring of health policies and the containment of disease progress and the registration of new cases. Research has suggested that there is a relationship between environmental issues and COVID-19 cases.

In this work, the results indicate that the population and demographic density (hab/km^2^) of a region are of great importance in the daily record of cases of COVID-19 infection and the distribution of cases, according to the age group, has not shown disparity between men and women.

Environmental factors such as RH%, average temperature °C, minimum temperature °C, maximum temperature °C, wind speed m / s and daily rainfall mm / day have a negative relationship with cases of COVID-19 in cities with humid equatorial climate in northern Brazil. In this region, a low wind speed and high RH are observed annually.

These environmental factors, when associated, reduce the virus transmission process because, they hinder the movement of the virus in the environment, which suggests that transmission in the region occurs directly through fluids between man /man and objects/man. Therefore, strategic public policies focused on controlling the pandemic must consider the environmental factors of the regions involved.

Studies show that room temperature and its correlation with RH are important factors in the transmission of the SARS-CoV-2 virus, as well as in mortality from CORVID-19 (Zhu et al., 2020; Ma et al., 2020). Therefore, it would be important to correlate these environmental factors in regions that have a low RH, high wind speed, high temperatures and low demographic density, to understand the behavior of these environmental factors in isolated transmission, when the population factor is not being relatively a factor critical.

Finally, attention is drawn to regions such as Peru (3rd), Ecuador (7th), Bolivia (12th), Honduras (13th) and Haiti (17th) on the rise in the ranking of COVID-19 cases in the Americas, as they are regions tropical climates very similar to the region of the present study. Especially for Haiti in Central America, as the region has a considerable demographic density of 361.5 inhabitants/km^2^, low Human Development Index (HDI = 0.404), life expectancy of 60 years. And Honduras, as it has a low RH and low temperature and moderate wind speed, these regions present meteorological combinations favorable to the spread of SARS-CoV-2, as identified in the results presented.

In South America, Ecuador has presented an increasing number of cases of COVID-19. This fact is worrying because it is a country with low temperatures, low Relative Humidity and mild wind speeds. These associated factors may favor the spread of SARS-Cov-2 in Ecuador.

## 4. Conclusion

The cases of COVID-19 in the RMB, have not shown disparities in relation to the gender/sex and age group of those infected. In terms of the relationship between environmental factors and the dynamics of daily cases, there is a low correlation between the factors RH%, average temperature °C, minimum temperature °C, maximum temperature °C, wind speed m/s and precipitation (rain) mm/day in number of COVID-19 cases.

However, the minimum temperature variable °C in the Santa Barbara region, showed a significant negative correlation at 5% with COVID-19 case records, which suggests a reduction in SARS-CoV-2 infections with increasing temperature. And therefore, in warmer periods in the region, it is expected that temperature is not a risk factor for infections.

Relative Humidity in the regions of Ananindeua and Castanhal showed a negative and relevant correlation with the cases of COVID-19, which suggests that RH is not a risk factor in the region, the spread of SARS-CoV-2, since the Relative Humidity recorded is always high.

It is noteworthy that in the northern region of Brazil the seasons of higher temperatures correspond to the period from June to December. As demonstrated in this study, high temperatures, despite the low negative correlation, were not statistically significant indicating that they reduce contamination and the registration of new daily cases. Likewise, there is no evidence to affirm that high temperatures are risk factors for the increase in cases of Covid-19. However, the significant negative correlation between number of cases and minimum temperature leads us to infer that high temperatures in the northern region will not favor the progress of SARS-CoV-2.

The multiple correlation of variables shows that high relative humidity associated with the phenomenon of precipitation, low wind speed and temperature above 26 ° C favors a lower incidence of new cases of COVID-19 in the study region. However, low relative humidity provides an increase in the number of cases and may be related to the drier environment. For, as the RH lowers, the environment becomes dry and facilitates the air transport of particles in the environment, enabling the transmission of the SARS-CoV-2 virus.

In the northern region, a low wind speed and high RH are observed annually. These associated environmental factors slow down the virus transmission process. Because, they hinder their movement in the environment, which suggests that the transmission of the virus in BMR occurs directly through fluids between man/man and objects/man.

Countries such as Haiti and Honduras in Central America, and Ecuador in South America are regions that must create and strengthen containment and public health policies aimed at combating pandemics and treating infected people, as they are regions that have favorable meteorological combinations for the spread of SARS -CoV-2, as identified in our results.

## Data Availability

All data are public in nature and available on the websites described below.

https://www.covid-19.pa.gov.br/#/

https://covid.saude.gov.br/

https://www.paho.org/bra/index.php?option=com_content&view=article&id=6101:covid19&Itemid=875

https://coronavirus.saude.gov.br/sobre-a-doenca#o-que-e-covid

https://coronavirus.saude.gov.br/

## Acknowledgments

This study was prepared with support from the Research Group on Biosystems Management, Modeling and Experimentation - GEMAbio. We thank Professor PhD. Mário Miguel Amin Garcia Herreros, for his commitment and dedication to education, contributions to GEMAbio research, the attention made available to students, colleagues and friends, and for his valuable and eternal friendship.

## References

Bashir, M. F., Ma, B., BilaL., Komal, B., Bashir, M. A., Tan, D., Bashir, M., Correlation between climate indicators and COVID-19 pandemic in New York, USA. 2020. Science of the Total Environment. v.728. 138835. DOI: https://doi.org/10.1016/j.scitotenv.2020.138835.

Bhattacharjee S., Statistical investigation of relationship between spread of coronavirus disease (COVID-19) and environmental factors based on study of four mostly affected places of China and five mostly affected places of Italy. 2020. arXiv preprint arXiv:2003.11277.

Bi, P., Wang, J., Hiller, J.E., 2007.Weather: driving force behind the transmission of severe acute respiratory syndrome in China? Intern. Med. J. 37 (8), 550–554. https://doi.org/10.1111/j.1445-5994.2007.01358.x.

BIGS. Brazilian Institute of Geography and Statistics. Populations. Available at: https://cidades.ibge.gov.br/brasil/pa/panorama. Date accessed: 10 May. 2020.

Brazil. Ministry of Health. Accumulated COVID-19 cases and deaths by confirmation date. Available at: https://coronavirus.saude.gov.br/sobre-a-doenca#o-que-e-covid. Access date: May 9 2020.

Bukhari, Q., Jameel, Y., 2020. Will Coronavirus Pandemic Diminish by Summer? 2020, SSRN Electronic Journal. DOI: http://dx.doi.org/10.2139/ssrn.3556998.

Chan, J.F.-W., Yuan, S., Kok, K.-H., To, K.K.-W., Chu, H., Yang, J., … Yuen, K.-Y., 2020. A familial cluster of pneumonia associated with the 2019 novel coronavirus indicating person-to-person transmission: a study of a family cluster. Lancet 395 (10223), 514–523. DOI: https://doi.org/10.1016/S0140-6736(20)30154-9.

Chatziprodromidou, T. A., and Vantarakis A. COVID-19 and Environmental factors. A PRISMA-compliant systematic review. medRxiv preprint DOI: https://doi.org/10.1101/2020.05.10.20069732.

Chatziprodromidou, T. A., and Vantarakis A. COVID-19 and Environmental factors. A PRISMA-compliant systematic review. 2020. medRxiv preprint DOI: https://doi.org/10.1101/2020.05.10.20069732.

Coccia, M. Factors determining the diffusion of COVID-19 and suggested strategy to prevent future accelerated viral infectivity similar to COVID. 2020, Science of the Total Environment, 729. DOI: https://doi.org/10.1016/j.scitotenv.2020.138474.

Hotez PJ, Bottazzi ME, Singh SK, Brindley PJ, Kamhawi S (2020) Will COVID-19 become the next neglected tropical disease? PLoS Negl Trop Dis 14(4): e0008271. https://doi.org/10.1371/journal.pntd.0008271

HSSP. Health Secretariat of the State of Pará. Coronavirus in Pará. Available at: https://portalarquivos.saude.gov.br/images/pdf/2020/May/09/2020-05-06-BEE15-Boletim-do-COE.pdf. Access date: 10 May. 2020.

Hulley S. B.; Cummings, S. R.; Browner, W. S.; Grady, D.; Hearst, N.; Newman, T. B. Delineando a pesquisa clínica: uma abordagem epidemiológica. 2a Ed. Porto Alegre: Editora Artmed; 2003.

Jahangiri, M., Jahangiri, Milad., Najafgholipour, M. The sensitivity and specificity analyses of ambient temperature and population size on the transmission rate of the novel coronavirus (COVID-19) in different provinces of Iran. 2020. Science of The Total. Environment. v.728, DOI. https://doi.org/10.1016/j.scitotenv.2020.138872.

Lal, P.; Kumar, A.; Kumar, S.; Kumari, S.; Saikia, P.; Dayanandan, A.; Adhikari, D.; Khan, M. L. The dark cloud with a silver lining: Assessing the impact of the SARS COVID-19 pandemic on the global environment. 2020, Science of the Environment, 732. DOI: https://doi.org/10.1016/j.scitotenv.2020.139297.

Lemaitre, J., Pasetto, D., Perez-Saez, J., et al., 2019. Rainfall as a driver of epidemic cholera: comparative model assessments of the effect of intra-seasonal precipitation events. Acta Trop. 190, 235–243. DOI: https://doi.org/10.1016/j.actatropica.2018.11.013.

Liu, j.; Zhou, J.; Yao, J.; Zhang, X.; Li, L.; Xu, X.; He, X.; Wang, B.; Fu, S.; Niu, T.; Yan, J.; Shi, Y.; Ren, X.; Niu, J.; Zhu, W.; Li, S.; Luo, B.; Zhang, k. Impact of meteorological factors on the COVID-19 transmission: A multicity study in China. 2020, Science of the Total Environment, 726. DOI: https://doi.org/10.1016/j.scitotenv.2020.138513.

Ma, Y., Zhao, Y., Liu, J., He, X.,Wang, B., Fu, S., et al., 2020. Effects of temperature variation and humidity on the death of COVID-19 in Wuhan, China. Sci. Total Environ. 724, 138226. DOI: https://doi.org/10.1016/j.scitotenv.2020.138226.

Menebo M. M. Temperature and precipitation associate with Covid-19 new daily cases: A correlation study between weather and Covid-19 pandemic in Oslo, Norway. 2020. Science of the Total Environment. v.737. 139659. https://doi.org/10.1016/j.scitotenv.2020.139659

Moraes, B. C.; Costa, J. M. N.; Costa, A. C. L.; Costa, M. H. Spatial and temporal variation of precipitation in the state of Pará. Acta Amazon, v. 35, p. 207–217, 2005.

Nikpouraghdam et al. Epidemiological characteristics of coronavirus disease (COVID-19) patients in IRAN: A single center study. 2020. Journal of Clinical Virology. 2019. DOI. https://doi.org/10.1016/j.jcv.2020.104378.

NMETI. National Meteorological Institute. Climatic data. Available and: http://www.inmet.gov.br/sim/sonabra/dspDadosCodigo.php?ODIxOTE=. Access date: May 07 2020.

Oliveiros, B., Caramelo, L., Ferreira, N.C., et al., 2020. Role of Temperature and humidity in the modulation of the doubling time of COVID-19 Cases. 2020, MedRxiv. DOI: https://doi.org/10.1101/2020.03.05.20031872.

ONU. World health statistics 2019: monitoring health for the SDGs, sustainable development goals. Geneva: World Health Organization; 2019. Licence: CC BY-NC-SA 3.0 IGO.

PAHO. Pan American Health Organization. Information sheet-COVID-19. Available in: https://www.paho.org/bra/index.php?option=com_content&view=article&id=6101:covid19&Itemid=875#datas-noticificacoes. access date: May 25 2020.

Prata, D. N., Rodrigues W., Bermejo, P. H. Temperature significantly changes COVID-19 transmission in (sub) tropical cities of Brazil. 2020. Science of the Total Environment. v.729. 138862. DOI. https://doi.org/10.1016/j.scitotenv.2020.138862.

şahin, M. Impact of weather on COVID-19 pandemic in Turkey. Science of the Total Environment. 2020. v.720. DOI. https://doi.org/10.1016/j.scitotenv.2020.138810.

Shi P., Dong Y., Yan H., Zhao C., Li X., Liu W., He M., Tang S., Xi S. 2020a. Impact of temperature on the dynamics of the COVID-19 outbreak in China. Science of the Total Environment. v.728, 138890. DOI: https://doi.org/10.1016/j.scitotenv.2020.138890.

Shi, P., Dong, Y., Yan, H., Li, X., Zhao, C., Liu, W., He, M., Tang, S., Xi, S., 2020b. The Impact of Temperature and Absolute Humidity on the Coronavirus Disease 2019 (COVID-19) Outbreak - Evidence from China. medRxiv. DOI: https://doi.org/10.1101/2020.03.22.200389192020.03.22.20038919.

Souza Filho, J. D. C.; Ribeiro, A.; Costa, M. H.; Cohen, J. C. P. 2005. Mechanisms to control the seasonal variation of transpiration in a tropical forest in northeastern Amazonia. Acta Amazônica 35: 223–229.

Tamerius, J.D., Shaman, J., Alonso, W.J., et al., 2013. Environmental predictors of seasonal influenza epidemics across temperate and tropical climates. PLoS Pathog. 9, e1003194. DOI: https://doi.org/10.1371/journal.ppat.1003194.

Tan, J.; Mu, L.; Huang, J.; et al., 2005. An initial investigation of the association between the SARS outbreak and weather:with the view of the environmental temperature and its variation. J. Epidemiol. Community Health 59, 186–192. DOI: https://doi.org/10.1136/jech.2004.020180.

Tosepu, R., Gunawan, J., Effendy, D.S., Lestari, H., Bahar, H., Asfian, P., 2020. Correlation between weather and Covid-19 pandemic in Jakarta, Indonesia. Sci. Total Environ. 725, 138436. DOI: https://doi.org/10.1016/j.scitotenv.2020.138436.

Van Doremalen, N.; Bushmaker, T.; Morris, D. H.; Holbrook, M. G.; Gamble, A.; Williamson, B. N.; Tamin, A.; Harcourt, J. L.; Thornburg, N. J.; Gerber, S. I.; Lloyd-Smith, J. O.; de Wit, E.; Munster, V. J. Aerosol and surface stability of SARS-CoV-2 as compared with SARS-CoV-1. 2020, N. Engl. J. Med. DOI: https://doi.org/10.1056.NJEMc2004979.

Wang, L., Li, J., Guo, S., Xie, N., Yao, L., Cao, Y., et al., 2020. Real-time estimation and prediction of mortality caused by COVID-19 with patient information based in algorithm. Sci. Total Environ. 138394. DOI: https://doi.org/10.1016/j.scitotenv.2020.138394.

World Health Organization. Coronavirus disease (COVID-19) outbreak situation. https://www.who.int/emergencies/diseases/novel-coronavirus-2019, Accessed date:15 April 2020.

Xie, J., Zhu, Y. Association between ambient temperature and covid-19 infection in 122 cities from China. 2020. Sci. Total Environ, 138201. DOI: https://doi.org/10.1016/j.scitotenv.2020.138201.

Zhou, P., Yang, X., Wang, X. et al.. A pneumonia outbreak associated with a new coronavirus of probable bat origin. 2020. Nature. v.579, p.270–273. DOI: http://doi.org/10.1038/s41586-020-2012-7.

Zhu, N., Zhang, D., Wang, W., et al., 2020. A novel coronavirus from patients with pneumonia in China, 2019. N. Engl. J. Med. DOI: https://doi.org/10.1056/NEJMoa2001017.

